# Single nuclei profiling identifies cell specific markers of skeletal muscle aging, sarcopenia and senescence

**DOI:** 10.1101/2021.01.22.21250336

**Authors:** Kevin Perez, Julia McGirr, Chandani Limbad, Ryosuke Doi, Joshua P Nederveen, Mats I Nilsson, Mark Tarnopolsky, Judith Campisi, Simon Melov

## Abstract

Aging is accompanied by a loss of muscle mass and function, termed sarcopenia, which causes numerous morbidities and economic burdens in human populations. Mechanisms implicated in age-related sarcopenia include inflammation, muscle stem cell depletion, mitochondrial dysfunction and loss of motor neurons, but whether there are key drivers of sarcopenia is not yet known. To gain deeper insights into age-related sarcopenia, we performed transcriptome profiling on lower limb muscle biopsies from 72 young, old and sarcopenic subjects using bulk RNA-seq (N = 72) and single-nuclei RNA-seq (N = 17). This combined approach revealed novel changes in gene expression that occur with age and sarcopenia in multiple cell types comprising mature skeletal muscle. Notably, we found increased expression of the genes *MYH8* and *PDK4*, and decreased expression of the gene *IGFN1*, in old muscle. We validated key genes in fixed human muscle tissue using digital spatial profiling. We also identified a small population of nuclei that express *CDKN1A*, present only in aged samples, consistent with p21-driven senescence in this subpopulation. Overall, our findings identify unique cellular subpopulations in aged and sarcopenic skeletal muscle, which will facilitate the development of new therapeutic strategies to combat age-related sarcopenia.

## Introduction

Age is the largest risk factor for developing sarcopenia -- a loss of skeletal muscle mass and function ^1^. Individuals over the age of 50 typically lose ~1% of muscle mass per year ^2,3^. Currently, physical activity appears to be the only therapy for sarcopenia, but its effectiveness declines with advanced age ^4,5^ and is only moderately successful. Loss of muscle mass can further cause frailty, and increase the likelihood of falls and fractures ^6^. The major mechanisms that drive age-related loss of muscle mass and function are unclear - although inflammation, impaired muscle regeneration due to loss of stem cells (satellite cells), loss of motor neurons and mitochondrial dysfunction have all been implicated ^7^.

To better understand potential mechanisms driving sarcopenia, several studies have examined changes in skeletal muscle gene expression with age. Differential expression with age has been reported for genes encoding proteins that participate in mitochondrial function, muscle structure and inflammation (e.g. mitochondrial ribosomal proteins, myosin heavy chain and IL-6, respectively). The majority of such studies used bulk RNA sequencing and modest sample sizes ^8,9^. Changes in muscle fiber types, notably a loss of type 2 fibers, is also a characteristic of aged skeletal muscle ^10^. Several aspects of the type 2 fiber gene expression profile can be reversed by exercise in conjunction with functional improvement ^11,12^.

At the cellular level, senescence is a cell fate that entails a stringent cell cycle arrest and a complex senescence-associated secretory phenotype (SASP) that can be induced by myriad stresses, including activated oncogenes, DNA damage, reactive oxygen species (ROS) and certain genotoxic chemotherapeutics ^13^. Senescent cells increase with age in many tissues, and are thought to contribute to sarcopenia ^14^. In addition, the SASP includes many pro-inflammatory factors that can cause chronic inflammation and alter tissue microenvironments to fuel the development of age-related diseases ^15^. Several markers of senescent cells have been identified, and include the cyclin-dependent kinase (CDK) inhibitors p16^INK4a^ (*CDKN2A*) and p21 (*CDKN1A*), which orchestrate proliferative arrest. However, there are no universally agreed upon drivers or biomarkers of senescence ^16^.

Single-cell analyses are improving our understanding of pathophysiology, including age-related diseases, by deconvolving the heterogeneity in cellular composition at the tissue level ^17^. Muscle “single cell” analyses have been particularly challenging to perform due to the nature of the myofiber: a syncytium containing thousands of myonuclei. This tissue organization practically prevents the preparation of “single cells” from muscle using conventional single cell workflows. Alternatively, nuclei from frozen tissue can be used to generate whole genome transcriptomes ^18-20^. This approach enables expression profiling of nuclei from muscle, despite its syncytial structure. Myonuclei within any muscle fiber express genes specific for different work capacities. Further, myofibers are broadly classified into two main types: type I (slow) and type II (fast) fibers. In addition to specific fiber types, skeletal muscle is also composed of numerous support cells, including endothelial cells, satellite cells, fibroadipogenic precursor cells and infiltrating immune cells ^21-23^. Bulk RNA-seq approaches can only provide a combined and aggregated view of the gene expression changes across all fiber and cell types.

Here, we contrast bulk RNA profiling of 72 biopsies from the lower limb muscles of young healthy, old healthy, and old sarcopenic human subjects, together with single nuclei sequencing (snucSeq) of 17 independent young and old muscle samples. Using the new technology of digital spatial profiling, which reports gene expression values at the genome wide level within the context of tissue microarchitecture ^24^, we also validate and localize several genes within individual fibers of young and old subjects. This combined approach provides a more complete insight into the heterogeneity of both cell composition and gene expression in aged individuals of differing functional capacities, and is generally applicable to other tissues. The methodology is also particularly useful for enumerating changes in cell frequency with age, a fundamental outcome of aging. Finally, we identified unique populations of cells in aged and sarcopenic muscle, including senescent cell types. Our results uncover potential new targets for therapies to treat age-related muscle loss and sarcopenia.

## Results

### Cohort description and clinical characteristics

We isolated 20-50 mg biopsies from the vastus lateralis of 72 adult men ^11,25^, including 19 young healthy subjects (avg. age = 20 years) and 53 older subjects (avg. age = 75 years). The older subjects were subsequently classified into healthy (N = 29) and sarcopenic (N = 24) subjects. Sarcopenia was diagnosed using multiple clinical factors that assessed both muscle strength and function [see Methods]. Older healthy subjects had lower short physical performance battery (SPPB) test scores (p < .05), longer ‘Timed up and Go’ performances scores (p < .05), lower grip strengths (p < .05), Biodex determined isometric knee extension torque (p < .05) and leg press scores (p < .05) relative to younger subjects. Sarcopenic subjects had even more impaired SPPB scores (p < .05), longer ‘Timed up and Go’ performances (p < .05), lower grip strengths (p < .05), isometric knee extension torque (p < .05) and leg press scores (p < .05) relative to older healthy subjects [Suppl. Figure 1, Suppl. Table 1].

### Changes in bulk gene expression with age and sarcopenia in human muscle

After quality control and processing using a standard RNA-seq pipeline [see Methods], we performed bulk RNA sequencing on biopsies from all 72 subjects. We then performed principal component analysis (PCA) on the resultant gene expression datasets [Figure 1A]. These analyses showed that gene expression at the bulk RNA level was largely distinct between younger and older subjects [Figure 1. A, Suppl. Data Table 1]. Samples from the older age group were marked by a statistically significant upregulation in the expression of several genes, including; *MYH8, COL19A1, EDA2R, CDKN1A* and *CDKN2B* [Figure 1B, 1C]. Conversely, expression of the following genes was lower with age: *IGFN1, MTND3P10, ATRNL1* and *PVALB*. Pathway analysis of the bulk data revealed several upregulated pathways consistent with increased inflammation with age [Figure 1D]. Sarcopenic and non-sarcopenic old muscle had similar gene expression profiles by these analyses, although the changes in sarcopenic subjects were of greater magnitude [Suppl. Figure 2A] (R^2^ = 0.9).

**Table 1.**
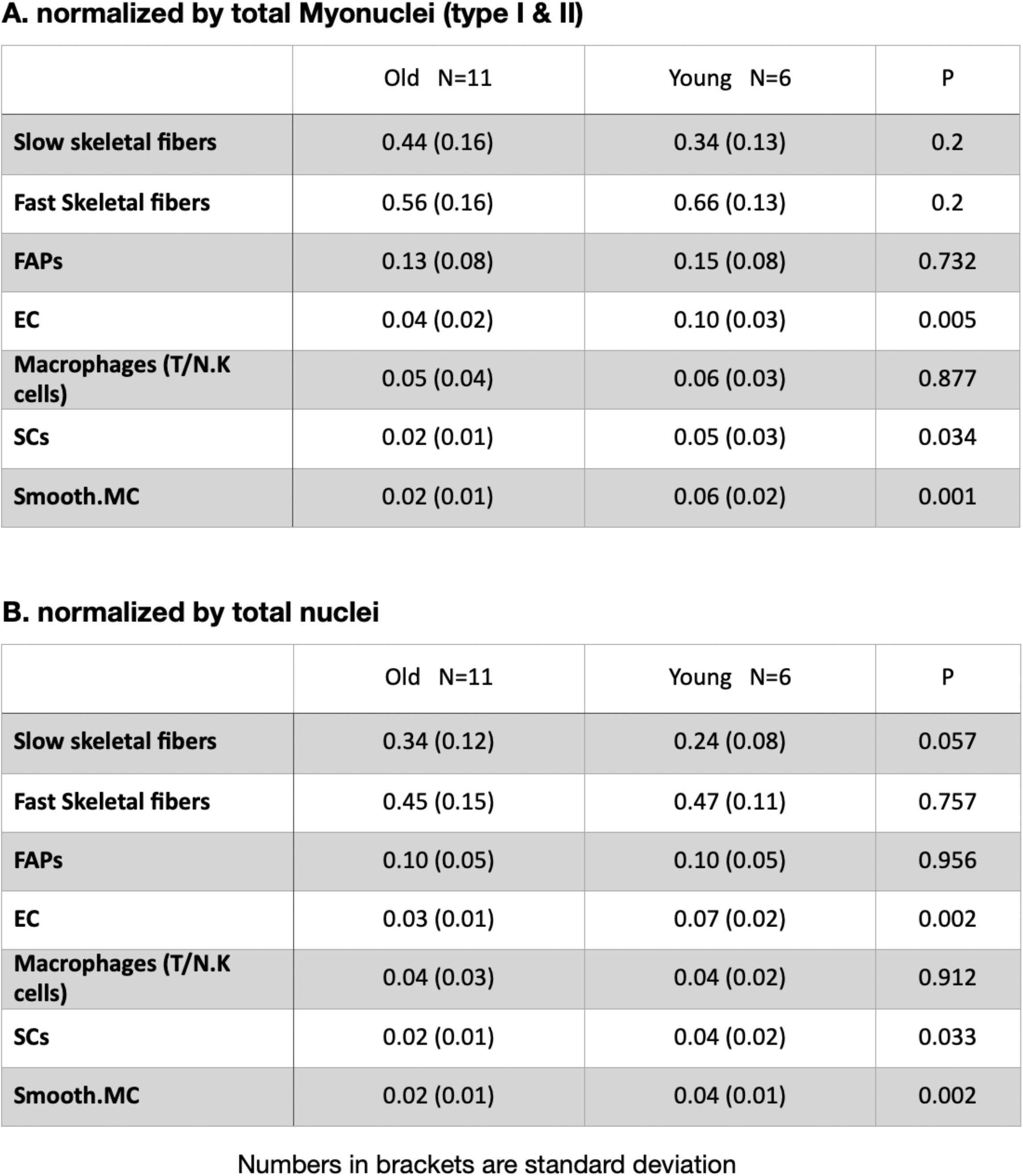
Proportions of cell types determined from nuclei in aged biopsies.

**A. normalized by total Myonuclei (type I & II)**
|  | Old N=11 | Young N=6 | P |
| --- | --- | --- | --- |
| **Slow skeletal fibers** | 0.44 (0.16) | 0.34 (0.13) | 0.2 |
| **Fast Skeletal fibers** | 0.56 (0.16) | 0.66 (0.13) | 0.2 |
| **FAPs** | 0.13 (0.08) | 0.15 (0.08) | 0.732 |
| **EC** | 0.04 (0.02) | 0.10 (0.03) | 0.005 |
| **Macrophages (T/N.K cells)** | 0.05 (0.04) | 0.06 (0.03) | 0.877 |
| **SCs** | 0.02 (0.01) | 0.05 (0.03) | 0.034 |
| **Smooth.MC** | 0.02 (0.01) | 0.06 (0.02) | 0.001 |

**B. normalized by total nuclei**
|  | Old N=11 | Young N=6 | P |
| --- | --- | --- | --- |
| **Slow skeletal fibers** | 0.34 (0.12) | 0.24 (0.08) | 0.057 |
| **Fast Skeletal fibers** | 0.45 (0.15) | 0.47 (0.11) | 0.757 |
| **FAPs** | 0.10 (0.05) | 0.10 (0.05) | 0.956 |
| **EC** | 0.03 (0.01) | 0.07 (0.02) | 0.002 |
| **Macrophages (T/N.K cells)** | 0.04 (0.03) | 0.04 (0.02) | 0.912 |
| **SCs** | 0.02 (0.01) | 0.04 (0.02) | 0.033 |
| **Smooth.MC** | 0.02 (0.01) | 0.04 (0.01) | 0.002 |

Numbers in brackets are standard deviation

**Figure 1.**
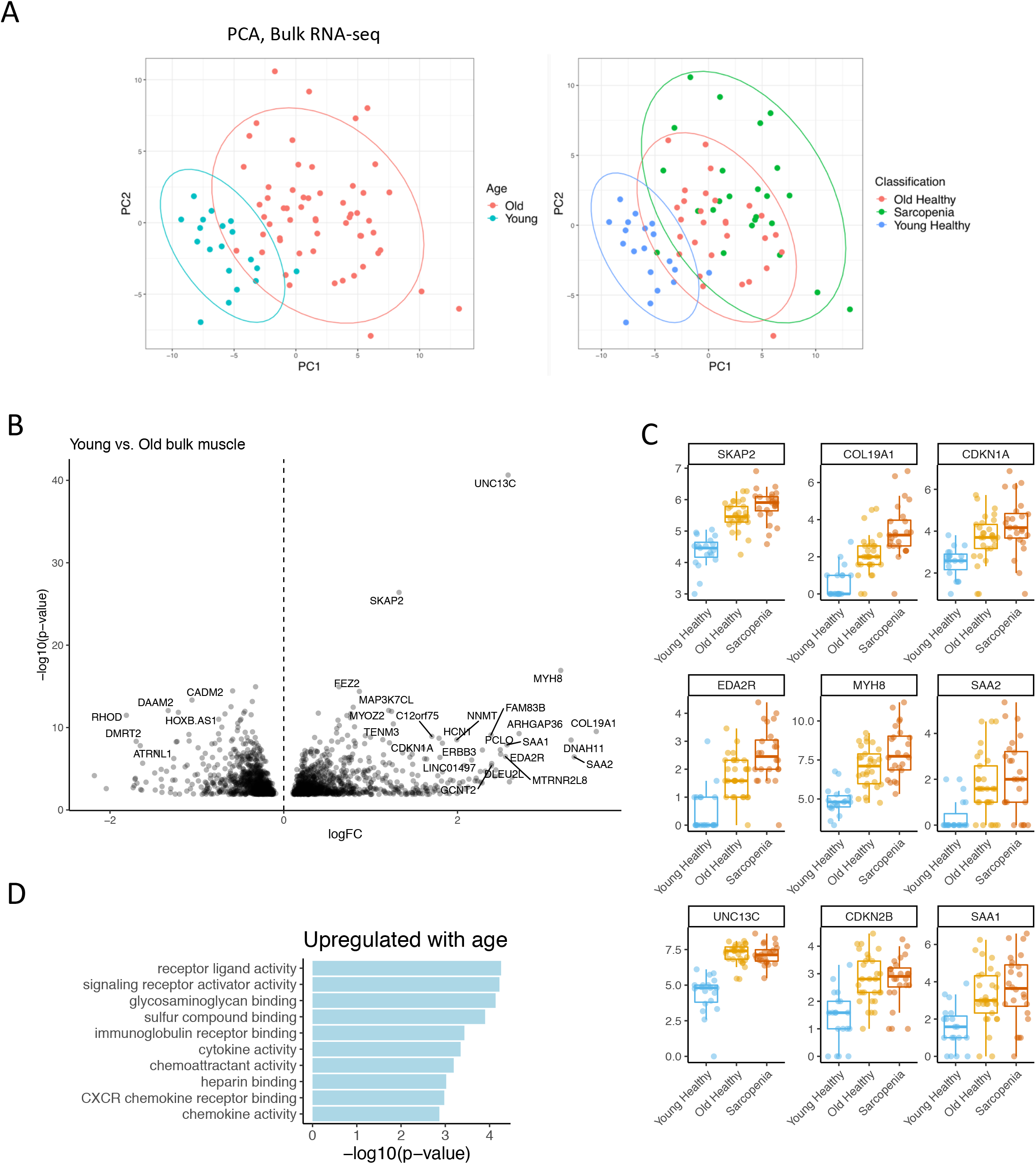
Bulk gene expression changes in muscle with age and sarcopenia. A. Principal component analysis (PCA) of bulk young, old and sarcopenic skeletal muscle. (left) Young (less than 20 yo) in blue, old (more than 65 yo) in red. (right) Young healthy (blue), old healthy (red), sarcopenic subjects (green). B. Volcano plot of expression changes in young vs old muscle. Labelled top 20 by abs(logFC)*–log10(p-value). C. Log(counts) of MYH8, COL19A1, MTRNL8, CDKN1A, CDKN2B, AREG in young healthy (green), old healthy (blue) and old sarcopenic subjects (red). Boxplot shows 25% percentile, 75% percentile and median. D. Pathway analysis of upregulated genes with age using KEGG, GO and Reactome database.

To determine whether the aging signature we observed in human skeletal muscle was conserved across mammalian species, we compared our results to a published multi-tissue rat aging gene expression study ^26^. Genes commonly upregulated in aged human and rat muscle included *CDKN1A, EDA2R, MUSK* and *CDKN2B* [Suppl. Figure 2B]. Comparing our muscle aging signatures with a published plasma proteome aging signature ^27^, we determined that CXCL11, EDA2R, MUSK and CXCL9 were upregulated with age in both muscle and blood [Suppl. Figure 2C].

### Genes associated with functional performance and age

We next asked whether these changes in gene expression were associated with clinical measures of health. Because age highly influenced most of the clinical metrics, we restricted this analysis to older subjects. Maintenance of muscle strength is generally considered more relevant to overall health at older ages ^28^. For each clinical metric, we classified the subjects as good performers (greater than the mean) or poor performers (lower than the mean). Leg press was the clinical metric associated with the most changes in gene expression, and grip strength was associated with the least changes. *RPL10P9* was upregulated in good performers of SPPB and ‘Timed up and Go’, whereas *PNPLA3* was downregulated in good performers of isometric knee extension torque and leg press [Suppl. Table 2]. Several genes were associated with both aging and muscle function. Indeed, one of the top downregulated genes with age - *IGFN1* -- was also upregulated in good performers of ‘Timed up and Go’. Similarly, *COL19A1*, one of the top upregulated genes with age, was downregulated in those with strong knee extensors (high isometric knee extension torque) [Suppl. Table 2]. These associations with both age and muscle function at older ages suggest a functional role for these genes in muscle decline, and therefore may serve as proxies for muscle function in the elderly.

### Single-nuclei sequencing reveals 7 clusters of unique cell types

To determine how cell composition changes with age in muscle, we analyzed 17 independent biopsies from the vastus lateralis of younger and older individuals (6 young, 11 old) using single nuclei sequencing (snuc-Seq). Single nuclei sequencing has several advantages in the context of tissues comprised of multiple cell types. Many single cell sequencing experiments rely on complex enzymatic digestion procedures to isolate cells of interest from the tissue. Isolation procedures can alter gene expression. Thus, tissue snap frozen at the time of isolation may best preserve the “in vivo” status of gene or protein expression. Hence, we employed a dedicated instrument for rapid extraction of nuclei from snap frozen fresh muscle tissue (Singulator, S2 Genomics) to rapidly isolate nuclei from the biopsies, thereby minimizing potential changes in gene expression resulting from conventional nuclei extraction procedures ^29^. We then carried out snuc-Seq using a 10x workflow to interrogate individual cell types in the skeletal muscle.

Following quality control, pre-processing and alignment, we generated 143,051 whole genome transcriptomes from individual nuclei (93,406 old, 49,645 young). After normalization and clustering, Uniform Manifold Approximation and Projection (UMAP) analyses revealed 7 distinct clusters, each corresponding to a unique cell type. We assigned cell identity to specific clusters using known markers for type II fibers, type I fibers, fibro-adipogenic progenitors (FAPs), satellite cells (SCs), smooth muscle cells, endothelial cells and immune cells (macrophages, T/NK cells) [Figure 2A, Suppl. Figure 3., Suppl. Figure 4., Suppl. Data Table 3]. After normalizing to total myonuclei (type I + II), we then calculated the proportion of type I (slow) and type II (fast) fibers per subject, which varied among the samples (Table 1). In an alternative normalization procedure, we determined the proportion of fiber types based on nuclei from all other cell types (Table 1). Regardless of normalization procedure, quantitation of fiber types within individuals agreed with values reported in the literature using histological staining (Table 1) ^30^. Similarly, we calculated the proportions of each major cell type between younger and older subjects. There was no significant difference in enumeration of nuclei associated with fiber types with age, FAPs, or macrophages. However, satellite cells, endothelial cells and smooth muscle cells were all significantly reduced with age (Table 1). Nuclei identified as derived from SCs in younger subjects comprised 5% of the nuclei population, consistent with prior reports, while older subjects had only 2% SC’s, consistent with reports of loss of this muscle specific stem cell with age ^31-33^.

**Figure 2.**
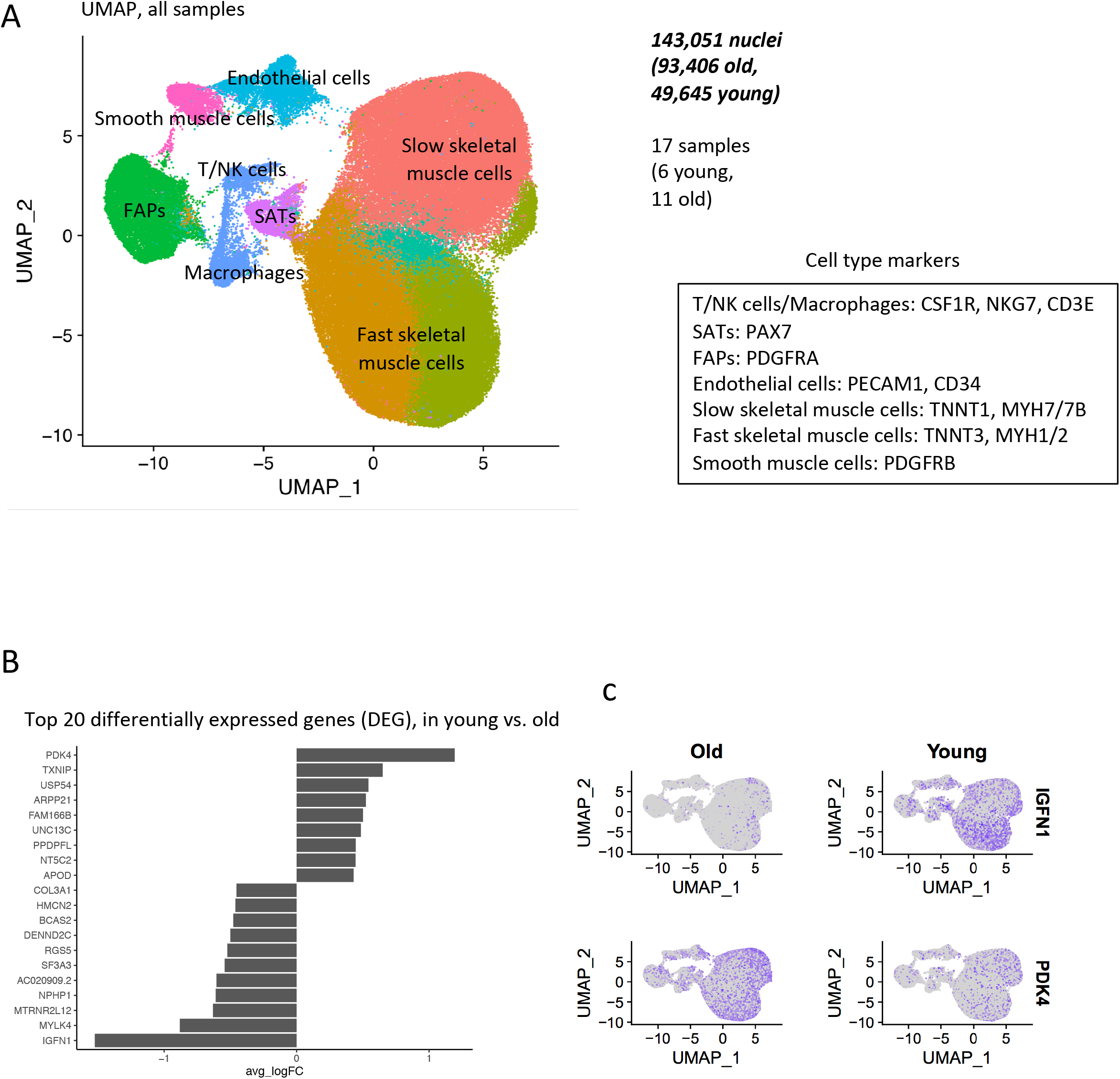
A. Uniform Manifold Approximation and Projection (UMAP) of 5’ single nuclei sequencing of human muscle. All samples are shown, after data normalization and Louvain clustering B. Top 20 differentially expressed genes (DEG), in young vs. old samples. All cells from all cell types are used in this test. Wilcoxon test, top 20 DEGs by logFC. C. Expression of PDK4 and IGFN1 in yound and old samples.

### *PDK4* is upregulated and *IGFN1* is downregulated with age in all cell types

We first examined broad changes between young and old samples by pooling all cell types. Notable were an upregulation of *PDK4* and downregulation of *IGFN1* expression. *IGFN1* was expressed in 49% of young sample nuclei, compared to 5% of old sample nuclei. *PDK4* was expressed in 15% of young nuclei, and 43% of old sample nuclei [Figure 2B, C]. We next compared the aging signatures obtained from the bulk study to this pooled single nucleus aging signature. Both bulk and single nuclei shared several commonly dysregulated genes, with *IGFN1* and *UNC13C* showing marked downregulation in both studies [Suppl. Figure 2. D].

We next examined cell-type specific changes with age. We identified 1,343 significant differentially expressed genes (DEGs), at a false discovery rate (FDR) of 1%. Some of these changes were common to many cell types, including *PDK4* and *IGFN1*, but most were cell type-specific. Of note, the transcriptomes that changed the most with age were from fast skeletal muscle fibers [Figure 3A]. The top 20 DEGs that changed by cell type with age [Figure 3B, Suppl. Data Table 2] included *PDK4, TXNIP*, HLA genes (*HLA-A, HLA-B, HLA-C, B2M*) and mitochondrial genes (*MT-ATP8, MT-CO3, MT-CYB). IGNF1, MTRNR2L12, MYLK4, NR4A1* declined in expression with age. It is well recognized that different muscle fiber types change dynamically with age. Among recognized fiber type markers ^34,35^, *MYH1* (fast type 2X), *MYH2* (fast type 2A), *MYH7* (slow type I fibers) and *MYH7B* (slow type I fibers) were present in our dataset, and *MYH2* expression significantly declined with age, consistent with type II fiber atrophy ^36^.

**Table 2.**
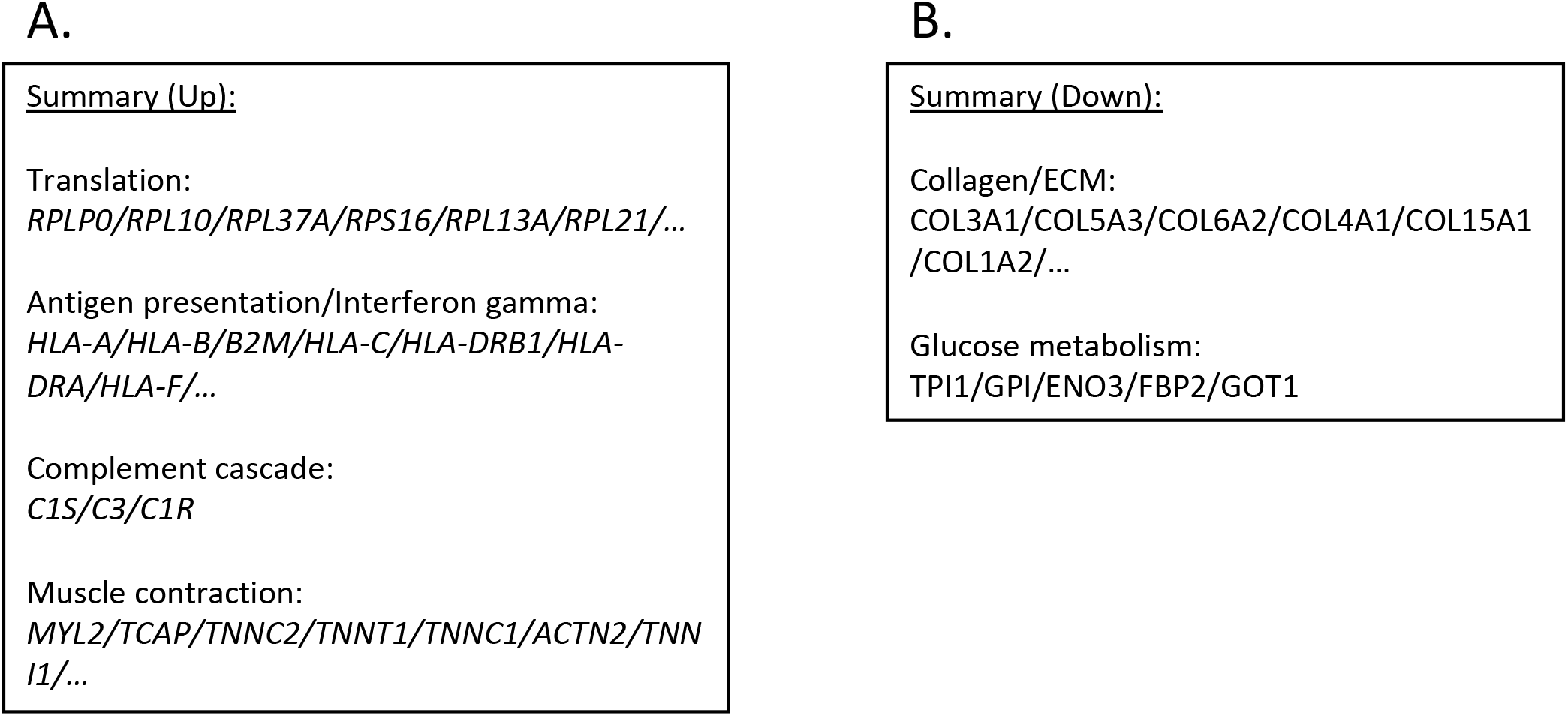
Summary of pathways dysregulated with age, including genes implicated in these pathways.

**Figure 3.**
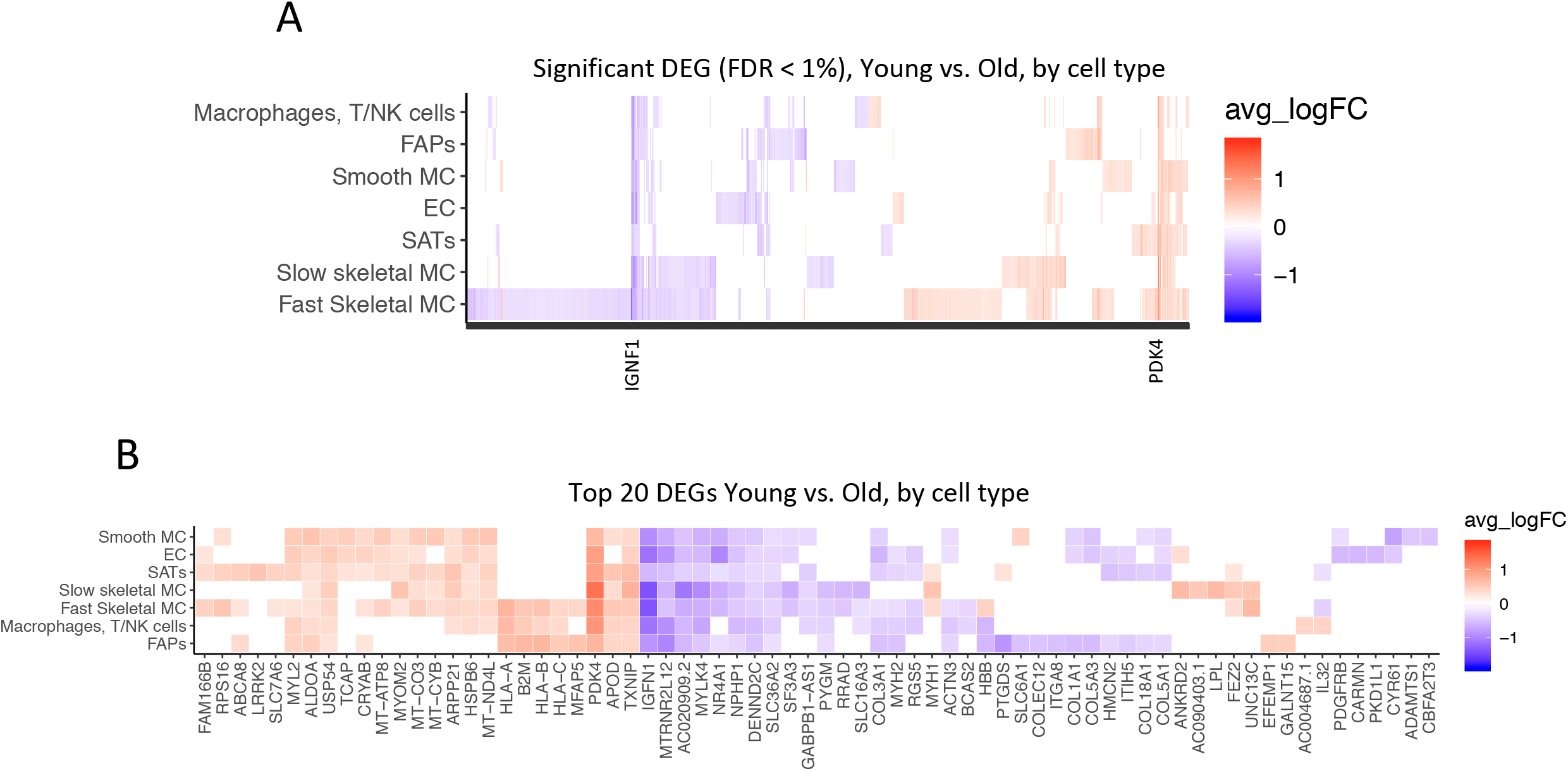
Changes with age by muscle cell type. Significant differentially expressed genes (DEG) in young versus older samples. A Wilcoxon test was performed for each gene in each cell type between samples, with a logFold-Change (logFC) threshold of .25, and False-Discovery Rate (FDR) < 1%. Red is upregulated with age, blue is downregulated. A. All DEGs are shown by cell type. B. Top 20 DEGs are shown by cell type, ranked by absolute logFC.

We performed a pathway analysis of the age-related changes within each cell type [Figure 4A, B]. Several pathways related to mRNA translation were upregulated with age in SMCs, SCs and fast SMC [Figure 4A]. Antigen presentation, gamma interferon responses and complement cascades were upregulated in immune cells [Figure 4A]. Muscle contraction pathways were upregulated in slow SMCs [Figure 4A]. Conversely, pathways related to collagen and extracellular matrix (ECM) declined with age in ECs, SCs, FAPs and SMCs [Figure 4B]. Glucose metabolism was downregulated in SMCs [Figure 4B]. Table 2 shows a summary of up and down regulated genes involved in the pathways.

**Figure 4.**
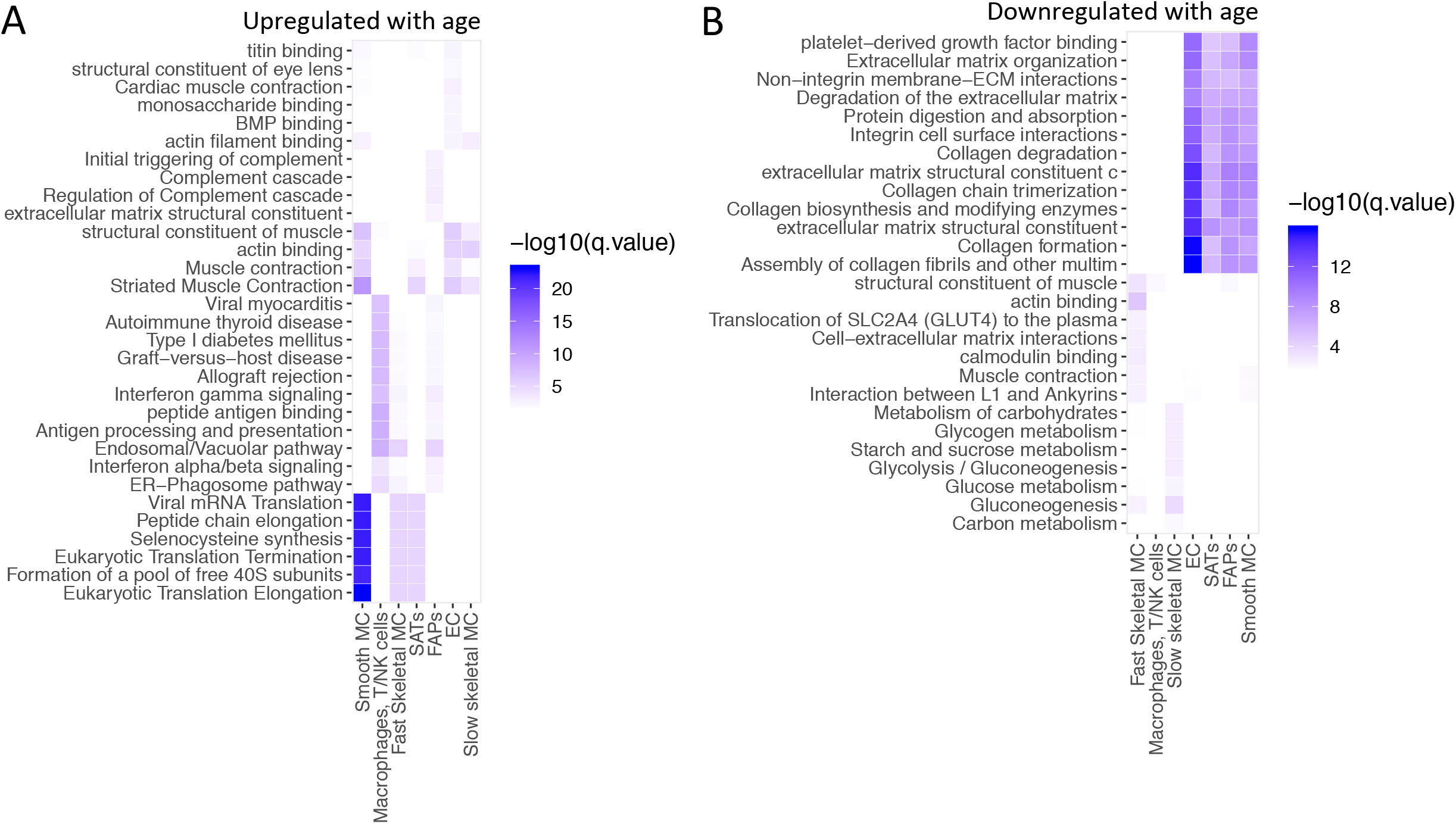
Pathway analysis of top 100 up-regulated and top 100 down-regulated genes with age in each cell type. GO, KEGG, Reactome pathways were queried. Over-representation was assessed using a hyper-geometric test at FDR 1%. A. Upregulated with age. B. Downregulated with age.

### Identification of a population of senescent cells in aged muscle

To investigate how cell composition in muscle changes with age, we determined the frequencies of different subtypes within each sample. Our approach bypasses laborious counting of histological sections to enumerate distinct cell types, or enzymatic digestions followed by isolation of cells with known specific markers using flow cytometry. It also has the capability to define novel cell subtypes that may drive pathology.

We first examined PDGFRA^+^ FAPs. We determined there are likely two main FAP subtypes – one expressed the muscle contraction gene Tryadin (*TRDN*), while the other expressed the skeletal muscle ryanodine receptor (*RYR1*) gene. Tryadin and RYR1 are known to interact, and are involved in muscle contraction ^37^. However, the frequency of neither subtype changed with age [Suppl. Figure. 5A]. We also identified infiltrating immune cells, including T/natural killer (NK) cells, macrophages and mast cells. There were no changes with age in the frequency of these immune cells [Suppl. Figure. 5B]. For CD34^+^ nuclei, we identified two main cell types. One cluster expressed *PDGFRB*, consistent with vascular smooth muscle cells. There was no change in the frequency of these cells with age [Suppl. Figure. 5C]. Consistent with prior reports, we observed an ~ 20-40% decrease in satellite cell frequency with age when normalized to total myonuclei (type I+II fibers) (Table 1). However, within SCs, which were Pax7^+^, we observed several distinct subclusters, possibly defining functional subgroups for this cell type. One subcluster expressed *LGR5* and *MYH7B*. Importantly, clusters 3 (*CDKN1A/MYH8/COL19A1/LRRK2*^+^) and 5 (*PDK4/TXNIP*^+^) significantly increased in frequency in samples from older subjects [Suppl. Figure. 6]. Lastly, we tested whether the transcriptomes derived from fast skeletal muscle fibers revealed more than one subtype. All clusters were present in equal proportions between younger and old samples, with one exception. This unique cluster contained *CDKN1A/MYH8/COL19A1/LRRK2*^+^ cells, and was present only in samples from older adults [Figure 5.]. Interestingly 3 of these genes (C*DKN1A, MYH8, COL19A1)* were among the top upregulated genes from the bulk study, suggesting that this small population could be responsible for the large variations we saw in the bulk study between younger and older age groups. Type II fibers are known to atrophy with age ^36^, but it remains to be determined the role of these specific nuclei in age-related atrophy.

**Figure 5.**
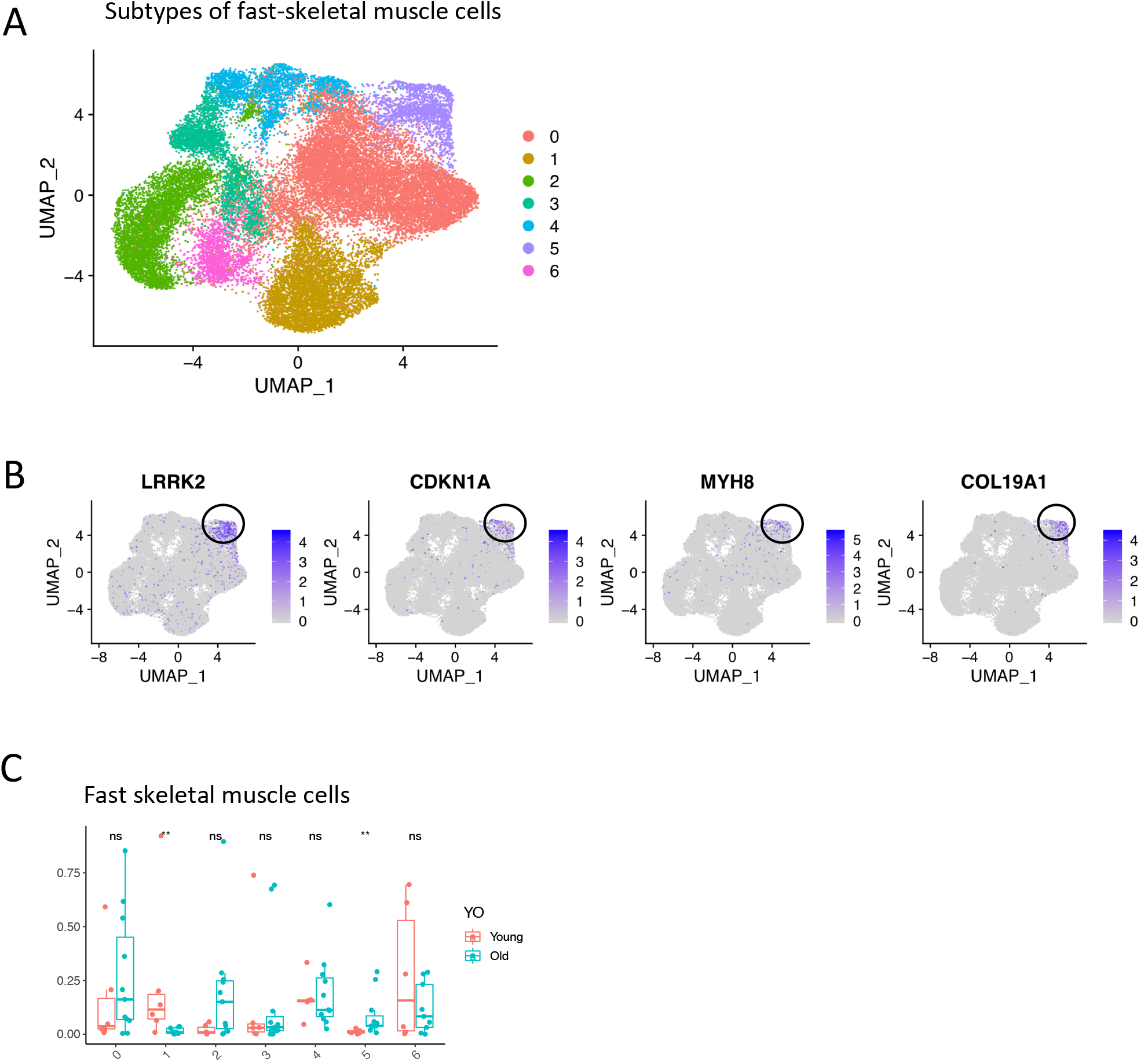
A. Subtypes of fast-skeletal muscle cells (UMAP, all samples). B. Cluster 5 is circled, with expression of LRRK2, CDKN1A, MYH8, COL19A1. C. Difference in proportions between young and old for all subtypes. Significance of the t-test between young and old is shown at the top of 5C.

### Spatial transcriptomics confirms gene expression determined from snuc-seq

To identify spatially resolved changes in gene expression between young and old muscle, we performed spatial transcriptomic profiling of two paraffin embedded human skeletal muscle biopsies (1 young, 1 old) using the GeoMx Digital Spatial Profiler (DSP, nanoString). Samples were stained with DAPI for nuclei and Desmin for muscle fibers [Figure 6A]. We then selected region of interest (ROIs) corresponding to longitudinal sections of muscle fibers, or, for some areas of the tissue section, cross-sections of muscle. Collectively, we determined whole genome profiles in tissue sections by DSP in 48 ROIs - 24 from young, 24 from old. After comparing gene expression in young and old fibers, old fibers had a distinct profile with upregulation of *PDK4, MT1X, MT2A* and downregulation of *IGFN1*, consistent with changes we identified in the single nuclei data [Figure 6B]. These changes were also consistent within individual ROI’s selected within the fibers [Figure 6C]. The majority of the fibers were type II (fast) fibers [Figure 6D].

**Figure 6.**
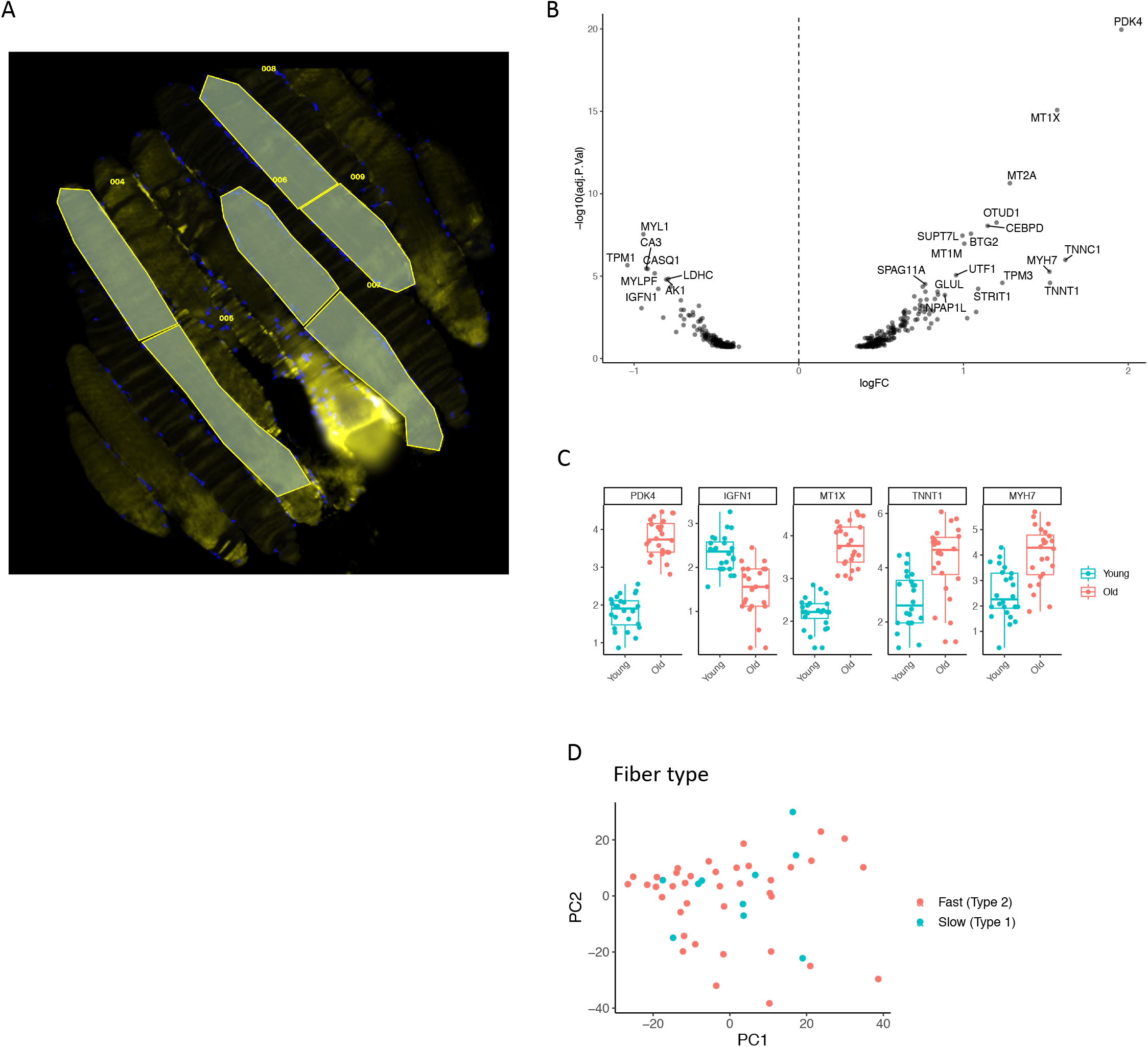
Spatial transcriptomics of young and old skeletal muscle. A. Young muscle fibers, several ROIs are shown in yellow delineating individual sections of distinct fibers. Myonuclei in blue, Desmin (muscle) in yellow. B Differentially expressed genes in young versus old spatial profiled muscle. C. log(counts) of top differentially expressed genes. E. PCA of fast and slow skeletal muscle types ROIs defined by type I/type II markers.

### CDKN1A, MYH8, COL19A1 and LRRK2 are upregulated in cultured senescent myoblasts

We further assessed whether the *CDKN1A*-expressing population of nuclei we detected in the old muscle samples was compatible with the expression profile of senescent myoblasts. We induced senescence in human primary myogenic cells (HSMMs), comprised of both myogenic progenitor cells and myotubes, using doxorubicin. After 7 days, we determined expression of the following genes: *CDKN1A, MYH8, COL19A1, LRRK2, EDA2R* and *PDK4* in senescent versus non-senescent cells by quantitative PCR (qPCR). In both cell types, all target genes were significantly upregulated in the senescent compared to non-senescent cells [Figure 7]. These results provide additional evidence that the expression profile of the population of cells we identified in aged skeletal muscle are likely senescent cells.

**Figure 7.**
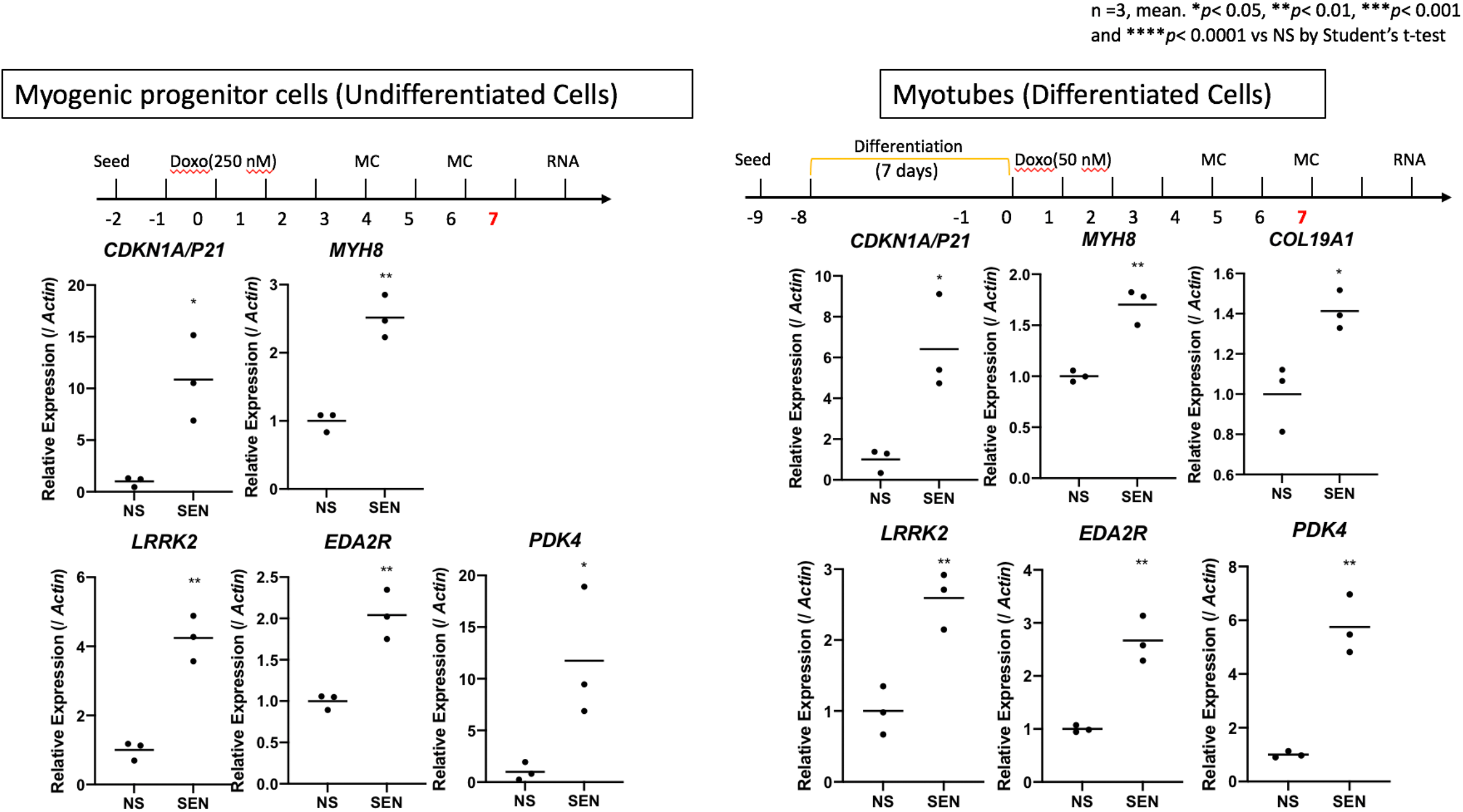
Quantitative PCR (qPCR) of CDKN1A, MYH8, COL19A1, LRRK2, EDA2R and PDK4 after 7 days of incubation in senescent vs. non-senescent cells. Senescence was induced using Doxorubicin in a cell line of Myogenic Progenitor cells (un-differentiated cells, left) and a cell line of Myotubes (differentiated cells, right). Expression is shown relative to Actin. 3 replicates in each condition/gene.

## Discussion

In this study, we identify changes in gene expression due to aging and sarcopenia in human skeletal muscle. We analyzed and verified gene expression signatures derived from bulk RNA-seq approaches and a more cell type-specific approach using single nuclei sequencing, digital spatial profiling, and cultured human skeletal muscle myoblasts.

At the bulk RNA-seq level, the top markers that increased with age and sarcopenia were *MYH8, COL19A1* and *CDKN1A*, in agreement with previous studies showing increased expression of *CDKN1A* (p21) and *MYH8* in muscle biopsies from older individuals ^12,38^. *MYH8* encodes the developmental protein myosin-heavy chain 8 (MYH8) ^35^. It is transiently expressed during embryonic development, but expression declines shortly after birth. It is also necessary for muscle regeneration after injury ^39^. The age-related rise in MYH8 expression is surprising, given its reported association with development and injury. Further, a *MYH8* mutation is associated with severe congenital muscle contractile disorders (distal arthrogryposis syndromes) ^35^. Thus, nuclei expressing MYH8 in aged skeletal muscle may indicate a response to age-related macromolecular damage, or may represent a unique marker of muscle dysfunction.

Interestingly, one prior report determined that HIV patients showed signs of premature muscle aging with similar gene expression signatures ^38^. Interestingly several markers that were upregulated in both rat and human muscle with age were senescent markers: cyclin-dependent kinase inhibitors CDKN2A (p16) and CDKN1A (p21), and the p53-dependent ectodysplasin A2 receptor (EDA2R) ^40,41^.

At the single nuclei level, we report broad upregulation of *PDK4* and downregulation of *IGNF1* expression in muscle from older individuals. *IGFN1* encodes the immunoglobulin like and fibronectin type III domain containing 1 protein (IGFN1), and is required for myoblast fusion and differentiation^42^. *PDK4* encodes pyruvate dehydrogenase kinase 4 (PDK4), and is overexpressed in skeletal muscle in patients with type 2 diabetes, fasting, and immediately post exercise ^43-46^. We also detected an upregulation of several HLA genes (*HLA-A, HLA-B, HLA-C*), including the HLA gene *B2M*, a putative pro-aging factor in plasma that could be detrimental for cognition ^47^. This finding is compatible with increased gamma interferon signaling with age ^48^, and support the long standing hypothesis that inflammation is a significant component of aging ^49^ and muscle dysfunction ^50^.

In support of prior reports ^10^, expression of *MYH2* (a type II fiber-specific gene) declined in older subjects. We also found a global decrease in the proportion of satellite cells in old muscle, consistent with a loss of muscle stem cells. Further, we identified a small population of *CDKN1A/MYH8/COL19A1/LRRK2*^+^ nuclei present only in muscle from older individuals. The expression profile of this population is consistent with that of senescent muscle cells, and may thus be targeted by senolytic compounds to improve muscle function ^51^. We also determined a number of previously discovered major aging pathways that were dysregulated in aged muscle. Collagen pathways declined with age, consistent with prior reports of decreased collagen production being a major phenotype of aging ^52^. Translation pathways also increased with age. Interestingly, TOR, a major regulator of aging, also regulates mRNA translation ^53^. Hence, our results reinforce existing literature implicating fibrosis and TOR dysregulation in aging skeletal muscle ^54,55^.

We found an association between differential gene expression changes at the bulk RNA level and clinical phenotypes. For example, *IGFN1* expression was associated with good performance (‘Timed up and Go’), and declined with age. Accordingly, we hypothesize that *IGFN1* has a beneficial role in aged muscle. In support of this idea, *IGFN1* mRNA was shown to increase in elderly muscle after an exercise training regimen ^56^. In contrast, low levels of *COL19A1* (Collagen XIX Alpha 1) expression were associated with good performance in the isometric knee extension torque (‘Biodex’), which may indicate that increased *COL19A1* expression relative to young individuals is detrimental to muscle function. High *COL19A1* expression also correlates with poor prognosis in Amyotrophic Lateral Sclerosis (ALS) ^57^.

Mature differentiated muscle is challenging to study due to the fact that muscle fibers are difficult to isolate, and many nuclei operate in concert along the fiber with yet to be elucidated zones of influence. One approach is to dissect out individual muscle fibers, and characterize them individually ^23^. An alternative approach is to use single-nuclei sequencing to capture gene expression from all nuclei in the fibers and all cells from a fresh muscle biopsy. In some cases, results from single nuclei can be concordant with bulk gene expression ^18^. The newly emerging technique of digital spatial profiling, for identifying gene expression within fixed or frozen tissue, offers great potential for localizing differentially expressed genes in the context of tissue micro-architecture. Here we applied this technique to young and old tissue sections of fixed skeletal muscle, and confirmed a number of differentially expressed genes with in situ localization to the messages within individual muscle fibers. Using our overall workflow, we were able to enumerate changes in frequency of all cell types with age, providing a new gold standard methodology for understanding cell composition changes with age.

Our study provides new insights into human skeletal muscle aging. It revealed remarkable heterogeneity in gene expression patterns in this tissue, both within individual fibers and between fibers and other cell types comprising skeletal muscle, and provided insights into mechanisms of muscle aging and sarcopenia. Using the array of technologies we describe here, we show for the first time that higher inflammation, lower mitochondrial capacity and stem cell exhaustion are likely *all* operative during aging and in sarcopenia. Our results suggest that broad approaches might be necessary in order to combat these age-related pathologies.

## Methods

### Ethics

All methods and procedures in this study were approved by the Hamilton Integrated Research Ethics Board (HIREB 2018-4656-GRA). Study participants were informed about potential risks of participation prior to giving informed consent. This study was part of a larger research project and collaboration between McMaster University (MAT), Exerkine Corporation (MAT), and Buck Institute for Research on Aging (Simon Melov, Novato, CA, USA) for evaluation of biomarkers of aging and muscle loss across different levels of sarcopenia. Specifically, some of the blood and muscle samples obtained at baseline from the participants were used for biomarker screening (muscle transcriptomics and serum proteomic analysis). Descriptive data and specific test outcomes, such as appendicular lean mass, maximal strength, and functional test results at baseline, were also shared between projects for characterization of subjects. This clinical trial was also registered at clinicaltrials.gov (NCT03536871).

### Subject recruitment

We recruited adults between the ages of 18-30 and 65-85 years old. Individuals were recruited from the community using advertisements in newspapers, at grocery stores, community centers and McMaster University. In addition, participants met with the study coordinators and had the trial explained to them.

### Inclusion Criteria

We recruited relatively inactive younger men and women (<1 hour of formal exercise/week) who were in the overweight category (body mass index or BMI 25 – 29.9 kg/m^2^). The healthy older male participants had a BMI of <30 kg/m^2^, muscle mass index of >7.23 kg/m^2^, and a 4-meter walk test of >0.8 m/s. Subjects in the older adult sarcopenia group had a BMI of <30 kg/m^2^, a muscle mass index of 8.51–10.75 kg/m^2^, and a 4-meter walk test of <0.8m/s.

### Exclusion Criteria

Medical conditions that precluded participation were diabetes mellitus (requiring more than one anti-diabetic drug), recent myocardial infarction (<6 months ago), hypertension (requiring more than two medications), congestive heart failure (requiring more than one medication), previous stroke with residual hemiparesis, renal disease (creatinine > 140), liver disease, musculoskeletal injury affecting exercise tolerance, musculoskeletal disorder (other than age-related SM wasting), severe osteoporosis, severe osteoarthritis, severe peripheral neuropathy, chronic obstructive pulmonary disease (FVC or FEV1 <70% of age-predicted mean value), asthma (requiring more than two medications), gastrointestinal disease, infectious disease, inability to consent, lactose intolerance/dairy protein allergy, and the use of medications affecting protein metabolism (for example, corticosteroids). Lifestyle-associated behaviors that precluded enrollment were smoking, veganism, recent weight loss or gain (<3-month period prior to the study), PA levels exceeding the minimal recommendations (150 min/week), and intake of supplements that affect musculoskeletal metabolism (e.g., whey, casein, calcium, creatine monohydrate, vitamin D, and omega-3 fatty acids).

### Subject classification

Subjects were classified into healthy young adults, healthy old adults and sarcopenic old adults according to the criteria in Suppl. Table 2.

### Biopsy collection

Participants arrived in the morning in a fasted state (10-12hr) and rested in the supine position for 10 minutes. A muscle biopsy was then taken from the vastus lateralis using local anesthetic as previously described ^25^.

### RNA extractions from muscle biopsies for bulk analysis

Muscle biopsies were homogenized using a mortar and pestle with liquid nitrogen. RNA was extracted from the powdered samples using the RNeasy Fibrous Tissue Mini Kit (Qiagen) and the QIAcube automatic processor (Qiagen). Integrity and concentration of the RNA was assessed using the Tapestation 4200 (Agilent Technologies), with the cut-off for acceptable integrity being an RNA integrity number (RIN) >7. Batch-tag-seq libraries were then produced from the RNA and run on the HiSeq 4000 by the DNA Technologies Core at U.C. Davis.

### Single nuclei RNA extraction and processing

Nuclei were isolated from the biopsies using the Singulator instrument (S2 Genomics). The instrument was primed with cold nuclei isolation and storage buffers (S2 Genomics) and the biopsy was loaded into the cartridge and covered with the 19.7 mm grinding cap. The “Nuclei_All_Tissues” protocol was used to isolate nuclei, after which the nuclei were centrifuged for 5 minutes and resuspended in buffer. The nuclei were counted using a Countess II Automated Cell Counter (Invitrogen), centrifuged again for 5 minutes and resuspended at the proper concentration for use in the Chromium Single Cell 5’ Library and Gel Bead Kit v1 (10X Genomics). Samples were processed using the Chromium Single Cell A Chip Kit and Chromium Controller (10X Genomics). Quality control was performed on the Tapestation 4200 (Agilent Technologies). Libraries were sequenced in one lane of a NovaSeqS4 by the U.C. Davis Genomics Core.

### Bulk RNAseq data analysis

Reads were aligned to the human genome using the STAR aligner and GRCh38 as the reference genome. Counts were computed using the featureCounts function in the subread software. Genes with a total count of less than 10 were removed from the analysis. PCA of differentially expressed genes were derived by the DESeq2 library in the R software, with absolute log Fold Change (logFC) > 1.5 and False-Discovery Rate (FDR) < 5%.

### Single-nuclei 5’ RNAseq data analysis

Reads were mapped to the human genome using Cell Ranger (10x Genomics), and the GRCh38 genome reference. Cells were removed if they expressed <200 unique genes. Genes not detected in any cell were removed from subsequent analysis. One sample was discarded due to low quality. Read count normalization, variable feature detection (nfeatures = 2000), scaling, UMAP (ndim = 10), and differential expression were computed as described in the Seurat package ^58^. Clustering was performed by the Louvain algorithm (resolution = 0.2). Cell types were characterized by a combination of known markers and de novo cluster markers [Table 1.].

### Pathway analysis

To derive the pathways containing differentially regulated genes, we performed a hyper-geometric test to assess over-representation. We used the clusterProfiler R package on the database Gene-Ontology (GO), Kyoto Encyclopedia of Genes and Genomes (KEGG), Reactome. The top 100 upregulated and downregulated genes were selected from the bulk RNA-seq study to execute the test. Pathways with p-value <0.05 were reported.

### Spatial transcriptomics

Two paraffin embedded human skeletal muscle biopsies (1 young, 1 old) were profiled using the GeoMx Digital Spatial Profiler (nanoSpring), as per the GeoMx protocol. Region of interests (ROIs) matching muscle fibers were selected in both biopsies. ROIs counts were normalized by area. Then, PCA, differential gene expression testing was performed using a linear model on each gene, comparing ROIs from young and old.

### Cell culture

Human skeletal muscle myoblasts (HSMMs; Lonza) were maintained at 37° C, 3% O_2_ and 5% CO_2_ in skeletal muscle growth media (SkGM2; Lonza). Differentiation was induced by replacing SkGM2 media with DMEM supplemented with 2% horse serum. For senescence induction, cells were treated with 50 nM (differentiated cells) or 250 nM (un-differentiated cells) of doxorubicin (Doxo) for 24 hours and then cultured for 7 days.

### qRT-PCR

Total RNA was isolated from HSMMs 7 days after Doxorubicin treatment using the RNeasy mini kit (Qiagen), and reverse-transcribed using the PrimeScript RT reagent kit (TAKARA Bio Inc.). Expression levels of the genes of interest were measured by real-time quantitative PCR using a CFX-384 instrument (BioRad). The sequences of the primer pairs are indicated in Table X1. The amount of mRNA was normalized to that of ACTIN mRNA.

### Primers

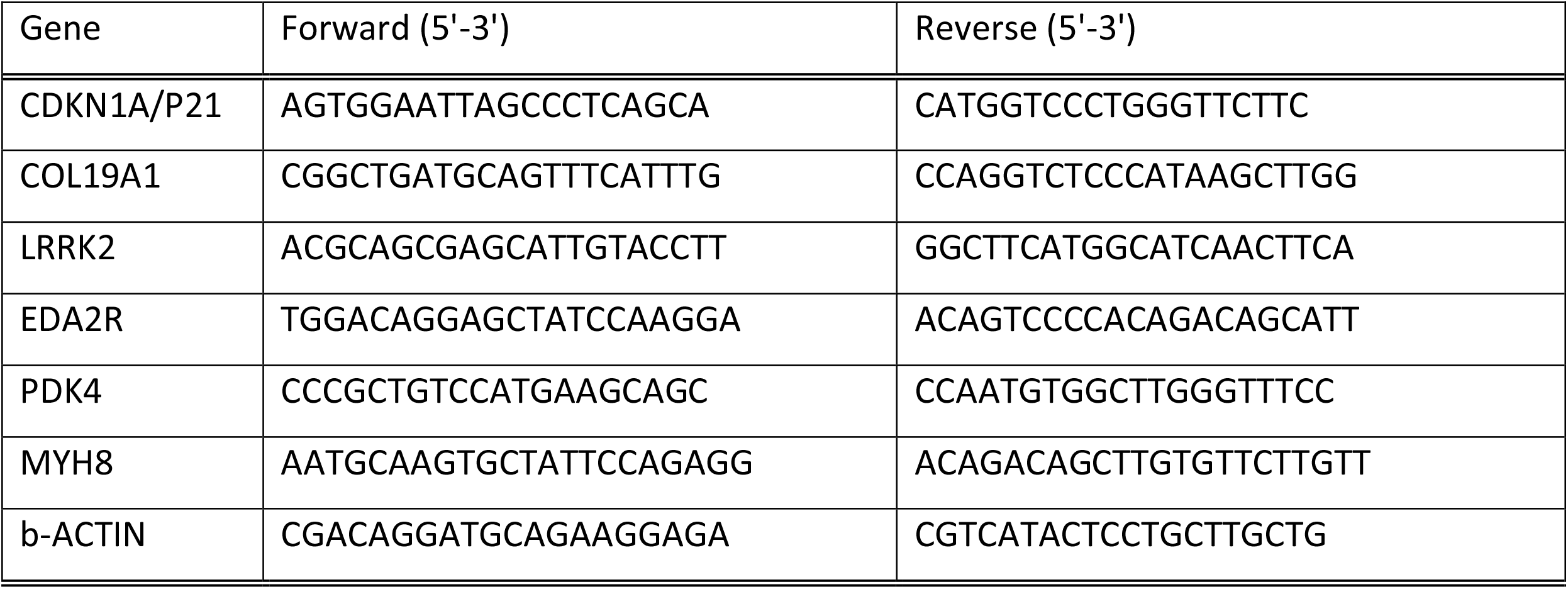

## Data Availability

Bulk and single cell RNA-seq counts and raw data have been posted on Gene Expression Omnibus (GEO), and will be released upon manuscript acceptance.

## Supplementary Methods

### Physical performance assessment

#### 1.1. Physical performance battery

A total of four functional tasks was performed once using a stopwatch that records to an accuracy of 0.01 second. For the 6-minute walk test, participants walked as fast as possible around a 20 m track for 6 minutes and distance measured to the closest meter. The standard 4-stair climb test involved participants climbing 4 stairs as fast as possible. The 5x sit-to-stand test involved participants performing a series of consecutive rising and sitting positions from a sturdy, armless plastic chair secured against a wall, with arms crossed at the chest. Finally, the chair rise-and-walk test involved starting from a seated position, standing and walking as quickly as possible in a predetermined straight line to a 9.14 m pylon, while going around the pylon, and returning to the original seated position.

#### 1.2. Leg press 1RM

Assessment procedures for determining lower body strength using leg press exercise equipment (Cybex Eagle ^®^, Medway, MA) required participant to sit in the leg press machine with the right and left foot on the weight platform. The seat and back pad were adjusted so that feet were flat on the platform a hip-width apart, toes slightly angled out and legs parallel to each other. The interviewer then instructed the participant to grasp the handles or sides of the seat and extend their legs leaving a slight bend in the knee. Next, the participant removed the racking mechanism from the platform and grasped the handles or seat again. The participant began with a selected weight that is within their perceived ability, ∼ 60 to 80% of maximum capacity. The participant lowered the platform slowly and controlled towards the chest, keeping hips and buttocks on the seat and the back flat against the back pad. Once the thighs were parallel to the platform, the participant extended the legs, pushing the weight back to the start position as hard and fast as possible. The participant was instructed to not allow hips to shift to one side, buttocks to rise or knees to move inward or outward during this exercise. The interviewer also instructed the participant to keep heels flat and not allow the knees to go beyond the toes. Once the repetitions were completed, the participant replaced the racking mechanism and exited the leg press. These procedures were adapted from those described by the National Strength and Condition Association (2008) and American College of Sports Medicine (2013).

#### 1.3. Hand grip strength (MVIC)

Hand grip strength was measured using an isometric dynamometer (JAMAR^®^, Sammons, Bolingbrook, IL). The grip width was adjusted to hand size, with the arm flexed at 90°. The participant performed three 5 s efforts with a one min rest between trials.

#### 1.4. Knee extension (MVIC)

Isometric knee extension was measured by mechanical dynamometry (Biodex System 3, Biodex Medical Systems, Shirley, NY). Participants were positioned in the machine with the knee flexed at 90° and performed 3 × 5s maximal voluntary contractions with 30s rests between each trial.

## Acknowledgements

We acknowledge the patients that contributed to this study. JPN was supported by a Canadian Institutes of Health Research (CIHR) Postdoctoral Fellowship. MAT was supported by a CIHR Foundation Grant (143325). Further support to JC and SM was from Astellas Pharmaceuticals, and NIH AG061879 (SM), AG051129(SM), and AG055822(SM & JC).

## Author Contributions

K.P. performed all data pre-processing and downstream analyses. J.M. performed the bulk RNA-seq and single-nuclei RNA-seq sample preparations. C.L. performed single-nuclei RNA-seq samples preparation. M.T. contributed to the design of the study and completed all of the muscle biopsies. J.P.N and M.I.N performed human testing and biopsy collection. S.M. designed the experiments and workflow, performed the spatial transcriptomics in conjunction with nanostring, and co-supervised the project with J.C. All authors contributed to writing and editing the manuscript.

## Data Availability

Bulk and single cell RNA-seq counts and raw data have been posted on Gene Expression Omnibus (GEO), (submitted).

---

**Suppl. Figure 1.**
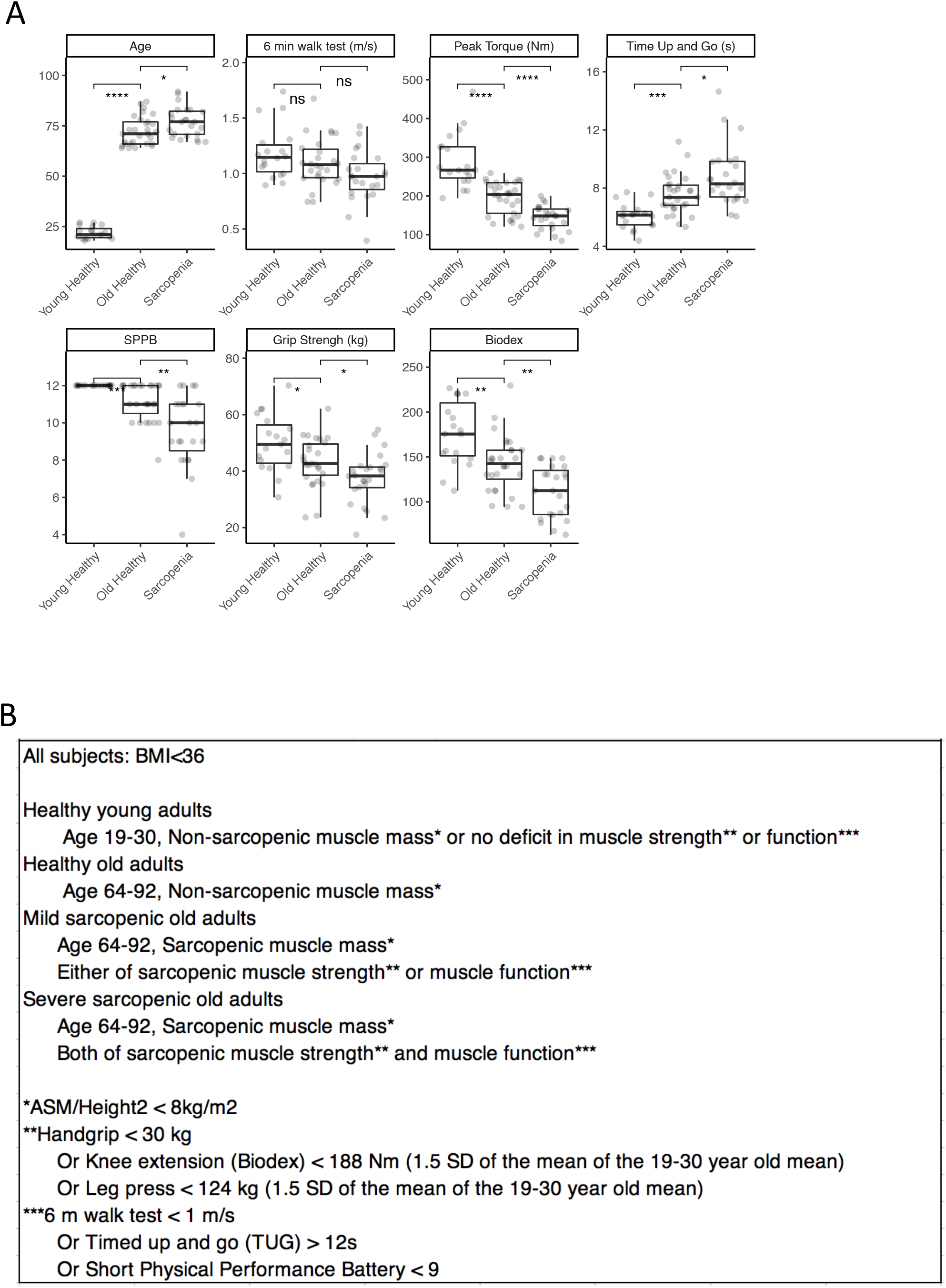
Clinical characteristics of the bulk cohort. Samples were classified as young healthy, old healthy, mild sarcopenic and severe sarcopenic using the criteria shown in B. In A., values for each group are shown with boxplots.

**Suppl. Figure. 2.**
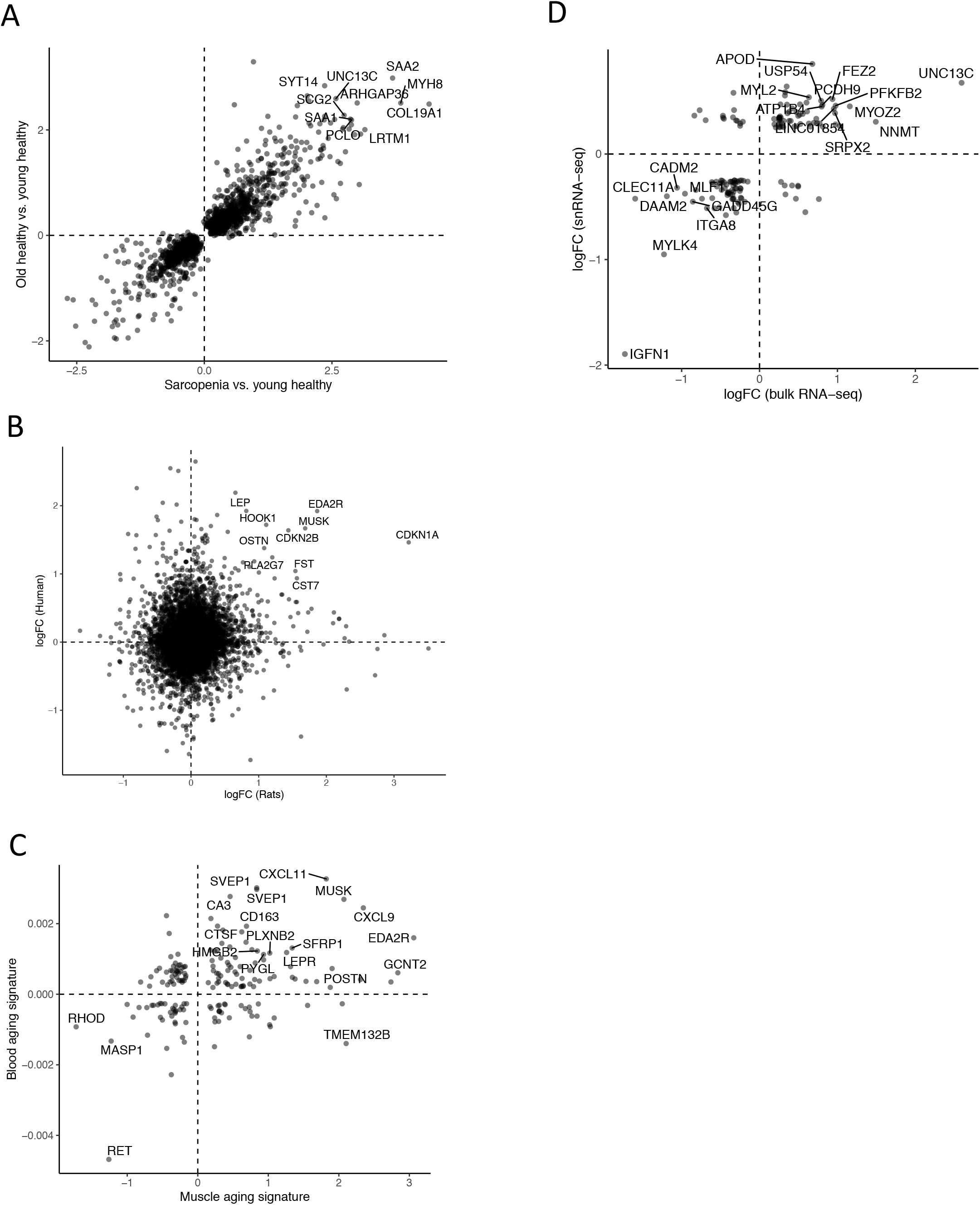
A. Sarcopenia vs. young healthy; Old healthy vs. young healthy (R^2^=0.9) B. Conserved aging signatures in rat and human C. Comparison of muscle and blood aging signature D. Comparison of bulk and single cell aging signatures All significantly differentially expressed genes (p-adj < .05) are shown. Labelled are top 20 genes with greatest logFC (X) * logFC (Y).

**Suppl. Figure 3.**
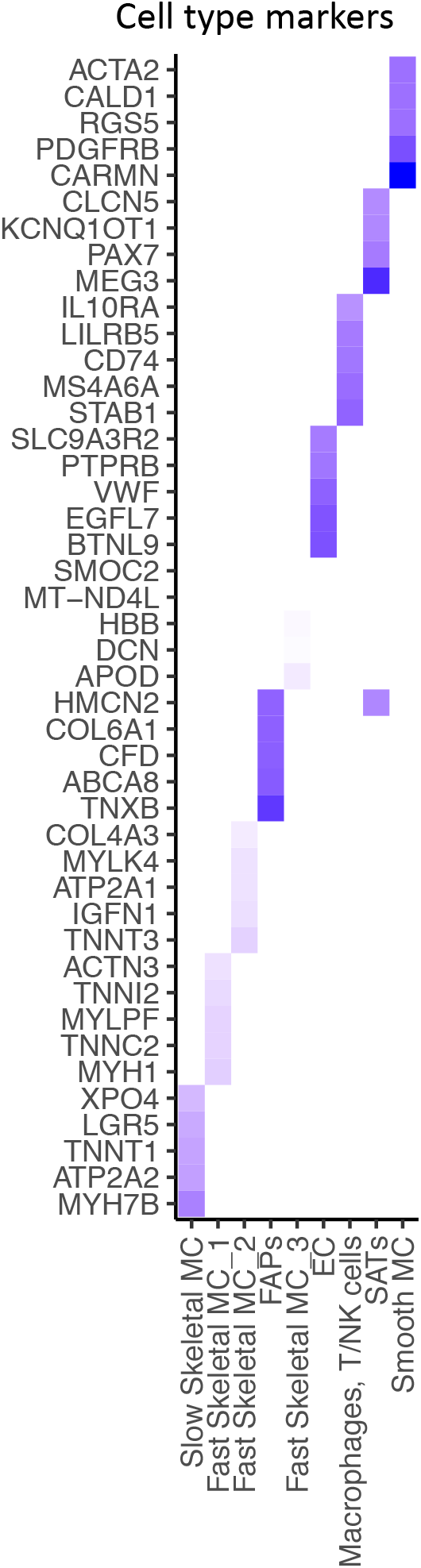
Cell type markers. Top 5 markers for each cell type, colored by logFC. B. Cell type specific pathways. Top 50 DEG per cell type were fed to the GO, KEGG, Reactome databases. Over-representation was assessed using an hyper-geometric test at FDR 1%.

**Suppl. Figure 4.**
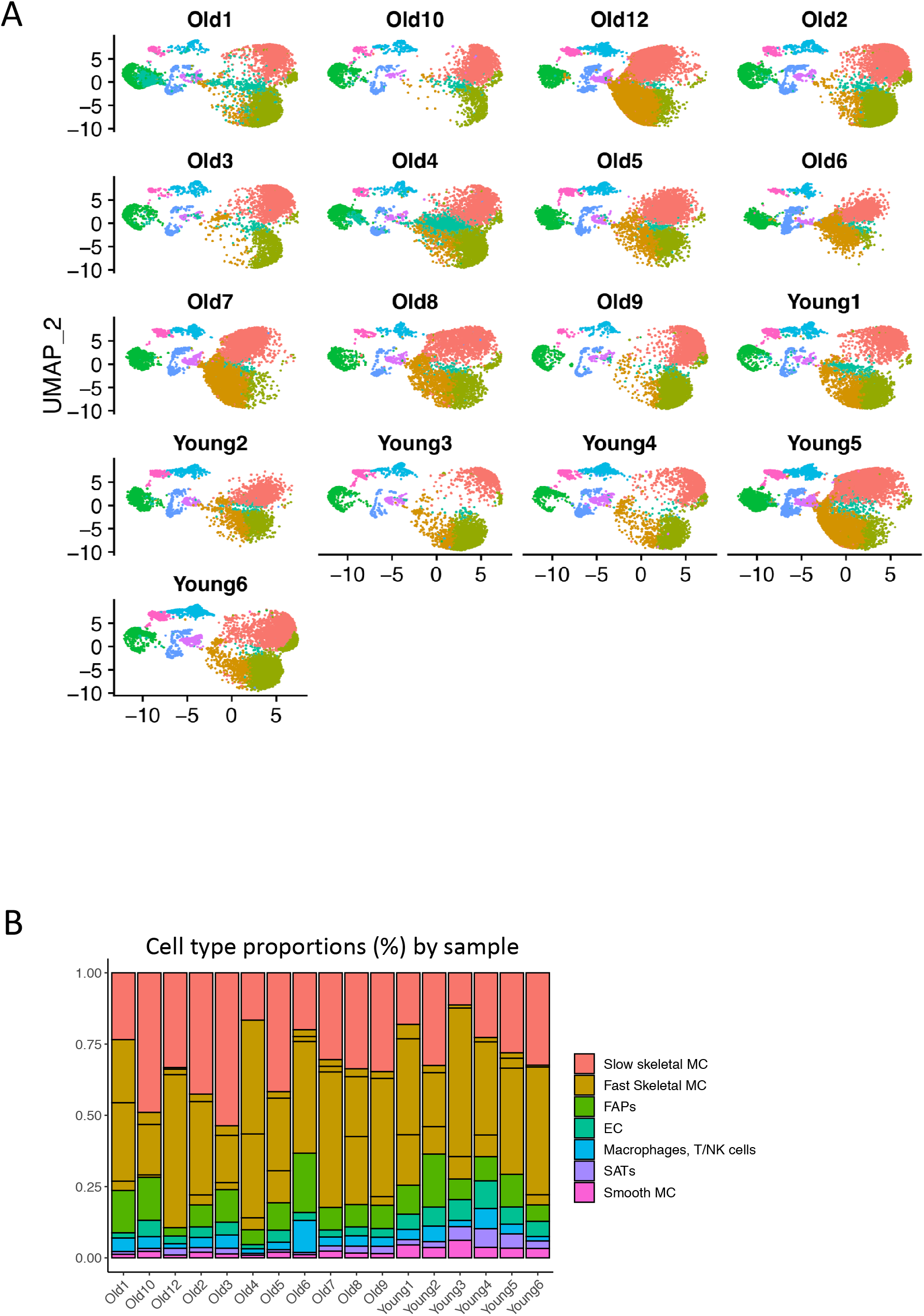
A. UMAP of each separate sample. B. Cell type proportions in each sample.

**Suppl. Figure 5.**
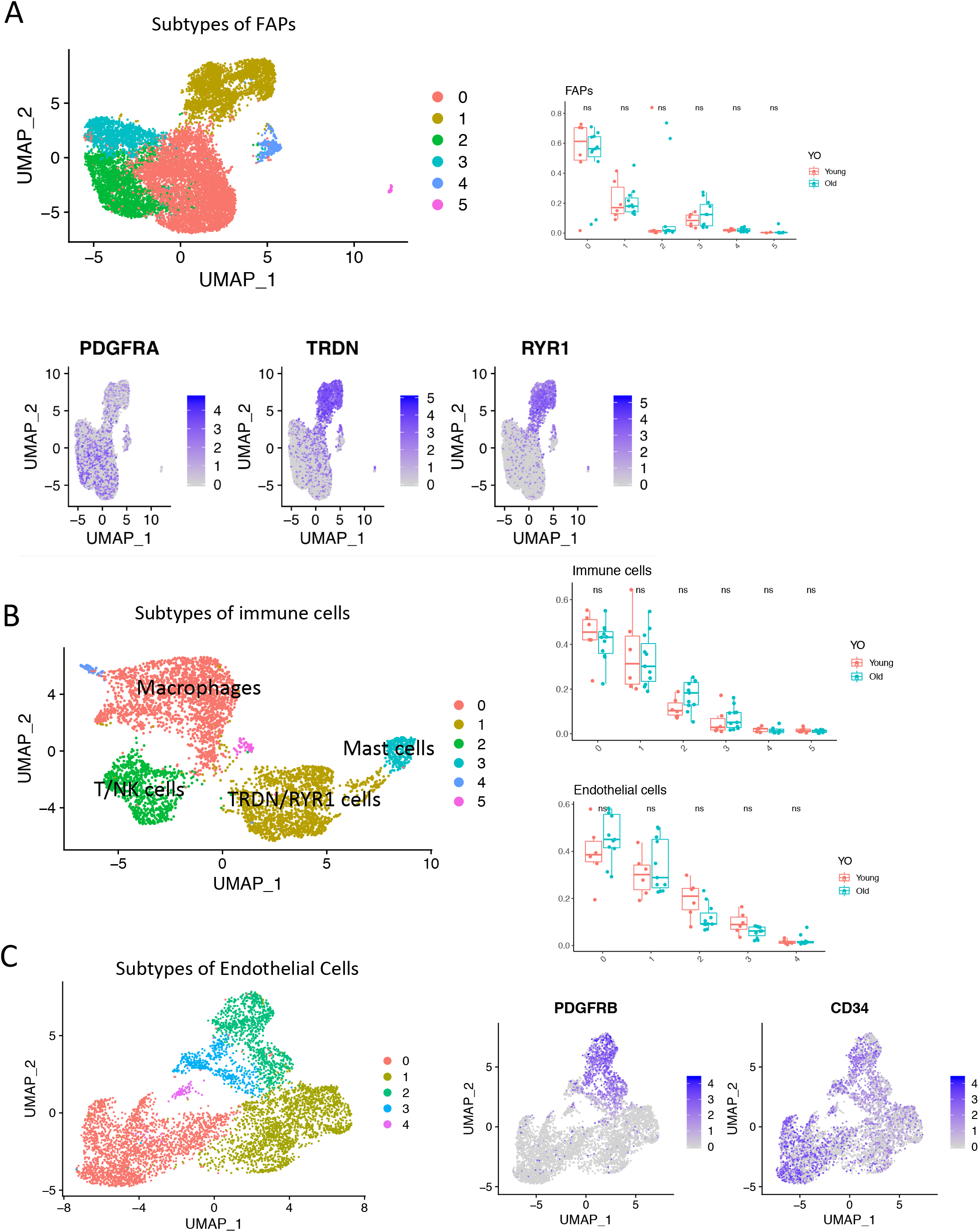
A. B. C. For the 3 cell types FAPs, immune cells, endothelial cells. Subtypes of each cell type is shown (UMAP, all samples). Markers expressed in different subtypes. Difference in proportions between young and old for all subtypes. Significance of the t-test between young and old is shown at the top.

**Suppl. Figure 6.**
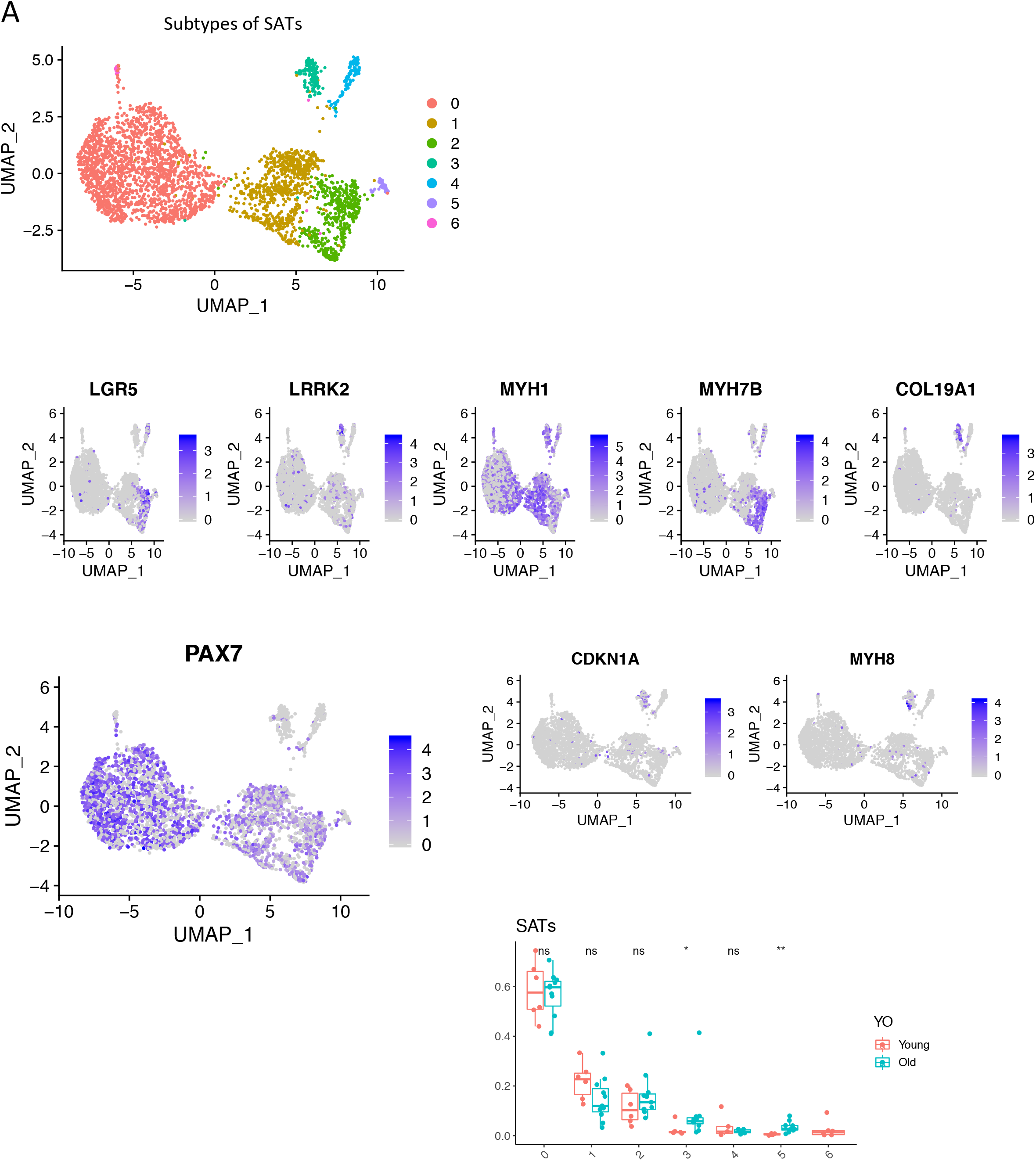
Subtypes of SATs (UMAP, all samples). Markers expressed in different cell types. Difference in proportions between young and old for all subtypes. Significance of the t-test between young and old is shown at the top.

**Suppl. Table 1.**
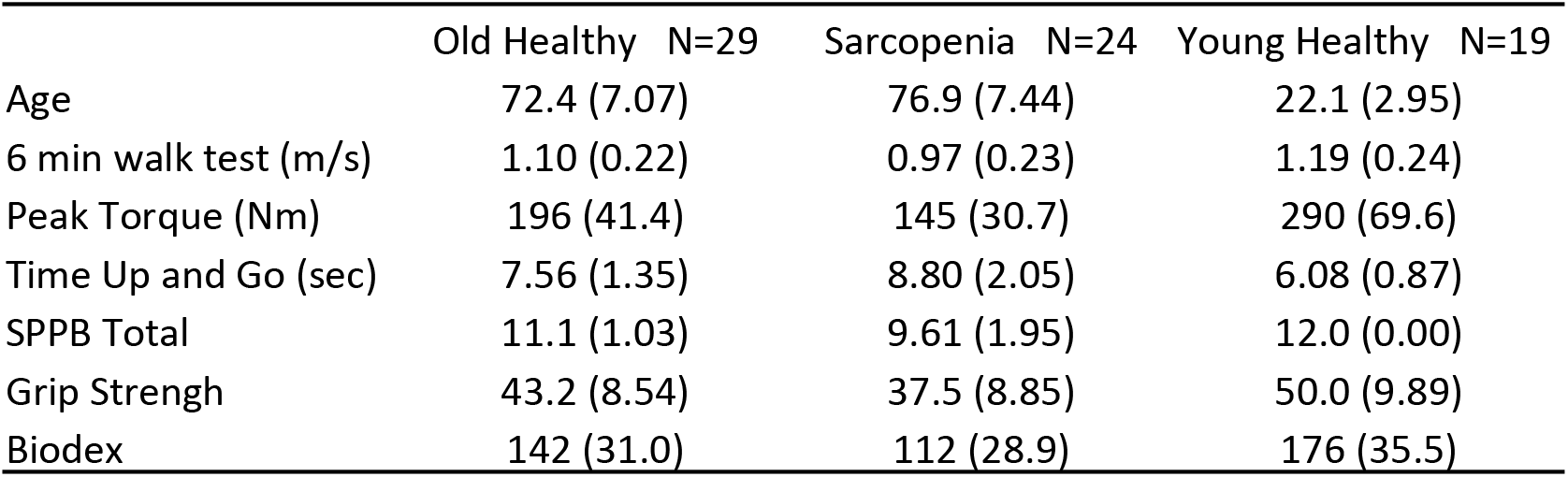
Mean (standard deviation) values for clinical parameters in the bulk cohort.

**Suppl. Table 2.** Assessment of changes in bulk RNA-seq with Clinical Parameters. Analysis of old (healthy & sarcopenic) samples only. Clinical factors were binarized to good and bad performers if they were above or below the median. DEGs between good and bad performers are shown for 6min walk test, SPPB, ‘Time up and Go’, grip strength, Biodex and leg press with P-value < .01, abs(logFC) > 2 are shown.

| Parameter | Up | Down |
| --- | --- | --- |
| **SPPB** | RPL10P9, CGA, MAP7D2 |  |
| **Grip Strength** | PPBP, SPAM1, SPATA17, LRRC65 |  |
| **Time Up and Go** | RPL10P9, GRP20, PPFIA3,<br>IGFN1, GAS2L2 | MTRNRL8, MTND4P24 |
| **6 min Walk Test** | MTCYBP35, PP2R2B, CDK18,<br>S100A2, CAMD5 |  |
| **Biodex** | GPR61, MAP7D2 | COL19A1, MYCL, LMO2, MPZL2,<br>PNPLA3, SLC47A2 |
| **Leg Press** |  | PAX5, COL25A1, NPTX1, PNPLA3 |

